# Application of MALDI-MS and Machine Learning to Detection of SARS-CoV-2 and non-SARS-CoV-2 Respiratory Infections

**DOI:** 10.1101/2023.08.31.23294891

**Authors:** Sergey Yegorov, Irina Kadyrova, Ilya Korshukov, Aidana Sultanbekova, Valentina Barkhanskaya, Tatiana Bashirova, Yerzhan Zhunusov, Yevgeniya Li, Viktoriya Parakhina, Svetlana Kolesnichenko, Yeldar Baiken, Bakhyt Matkarimov, Dmitriy Vazenmiller, Matthew S. Miller, Gonzalo H. Hortelano, Anar Turmuhambetova, Antonella E. Chesca, Dmitriy Babenko

**Affiliations:** Michael G. DeGroote Institute for Infectious Disease Research; McMaster Immunology Research Centre; Department of Biochemistry and Biomedical Sciences, McMaster University, Hamilton, ON, Canada; School of Sciences and Humanities, Nazarbayev University, Astana, Kazakhstan; Research Centre, Karaganda Medical University, Karaganda, Kazakhstan; National Laboratory Astana, Centre for Life Sciences, Nazarbayev University, Astana, Kazakhstan; School of Engineering and Digital Sciences, Nazarbayev University, Astana, Kazakhstan; City Centre for Primary Medical and Sanitary Care, Karaganda, Kazakhstan; Infectious Disease Centre of the Karaganda Regional Clinical Hospital, Karaganda, Kazakhstan; Department of Internal Diseases, Karaganda Medical University, Kazakhstan; Faculty of Medicine, Transilvania University, Braşov, Romania

**Author notes:** **Corresponding authors’ contact emails:** (SY), (IK).

**Keywords:** Acute respiratory infection, COVID-19, SARS-CoV-2, MALDI-MS, Machine Learning

## Abstract

**Background:** Matrix-assisted laser desorption/ionization mass spectrometry (MALDI-MS) could aid the diagnosis of acute respiratory infections (ARI) owing to its affordability and high-throughput capacity. MALDI-MS has been proposed for use on commonly available respiratory samples, without specialized sample preparation, making this technology especially attractive for implementation in low-resource regions. Here, we assessed the utility of MALDI-MS in differentiating SARS-CoV-2 versus non-COVID acute respiratory infections (NCARI) in a clinical lab setting of Kazakhstan.

**Methods:** Nasopharyngeal swabs were collected from in- and outpatients with respiratory symptoms and from asymptomatic controls (AC) in 2020-2022. PCR was used to differentiate SARS-CoV-2+ and NCARI cases. MALDI-MS spectra were obtained for a total of 252 samples (115 SARS-CoV-2+, 98 NCARI and 39 AC) without specialized sample preparation. In our first sub-analysis, we followed a published protocol for peak preprocessing and Machine Learning (ML), trained on publicly available spectra from South American SARS-CoV-2+ and NCARI samples. In our second sub-analysis, we trained ML models on a peak intensity matrix representative of both South American (SA) and Kazakhstan (Kaz) samples.

**Results:** Applying the established MALDI-MS pipeline ”as is” resulted in a high detection rate for SARS-CoV-2+ samples (91.0%), but low accuracy for NCARI (48.0%) and AC (67.0%) by the top-performing random forest model. After re-training of the ML algorithms on the SA-Kaz peak intensity matrix, the accuracy of detection by the top-performing Support Vector Machine with radial basis function kernel model was at 88.0, 95.0 and 78% for the Kazakhstan SARS-CoV-2+, NCARI, and AC subjects, respectively with a SARS-CoV-2 vs. rest ROC AUC of 0.983 [0.958, 0.987]; a high differentiation accuracy was maintained for the South American SARS-CoV-2 and NCARI.

**Conclusions:** MALDI-MS/ML is a feasible approach for the differentiation of ARI without a specialized sample preparation. The implementation of MALDI-MS/ML in a real clinical lab setting will necessitate continuous optimization to keep up with the rapidly evolving landscape of ARI.

## Introduction

The global response to the COVID-19 pandemic has underscored gaps existing in the laboratory-based diagnosis of acute respiratory infection (ARI)(1). In the early stages of the pandemic, a shortage of rapid and inexpensive techniques amenable to modification to adapt to the newly characterized SARS-CoV-2 motivated the search for alternative diagnostic tools. Matrix-assisted laser desorption/ionization mass spectrometry (MALDI-MS), a technique traditionally employed in proteomics and metabolomics, has emerged as a promising alternative to molecular and immunochromatography-based assays to detect SARS-CoV-2 (2). Several different MALDI-MS-based approaches involving varied degrees of sample preparation have been described (2).

Our clinical laboratory has particularly been interested in the”untargeted” MALDI-MS method, which applies Machine Learning (ML) algorithms to discern SARS-CoV-2 infection using MALDI-MS peak matrices acquired from respiratory samples such as nasopharyngeal swabs (NPS) without a specialized sample preparation (3–5). Therefore, in this study, we explored the feasibility and accuracy of such untargeted MALDI-MS/ML in differentiating SARS-CoV-2 from non-COVID acute respiratory infections (NCARI) in a clinical laboratory setting in Kazakhstan.

## Materials and Methods

### Study setting

We collected NPS from three participant subgroups: symptomatic SARS-CoV-2+, NCARI and asymptomatic controls (AC). Participants were recruited between May 25, 2020, and December 20, 2022. Written consent was obtained from all adult participants in the presence of a study coordinator; parental consent was obtained for participants under 18 years of age. The ARI diagnosis was made based on the presence of at least one of the following: fever, nasal congestion, cough, sore throat, and/or lymphadenopathy. SARS-CoV-2+ participants were recruited from among patients of the Karaganda regional clinical hospital, hospitalized with a PCR-confirmed SARS-CoV-2 infection. NCARI participants were recruited at the Karaganda regional clinical hospital and the Karaganda City Centre for Primary Healthcare among patients admitted for moderate-severe ARI symptoms. Most (72.4%) NCARI participants were PCR-positive for common respiratory viruses (adenovirus, seasonal coronaviruses, bocavirus, parainfluenza viruses, respiratory syncytial virus, rhinovirus, influenza or metapneumovirus) or bacteria (*Chlamydia pneumoniae* or *Mycoplasma pneumoniae)*. Samples were collected around day 3 (median, IQR [2-4]) and day 5 (median, IQR [3-7]) post-symptom onset for the SARS-CoV-2+ and NCARI participants, respectively. The AC sub-group was recruited from amidst the Karaganda University employees. The SARS-CoV-2 infection status was confirmed in the research lab for all samples using SARS-CoV-2 PCR as described earlier [6, 7]. All samples were frozen at -80C until processing.

In addition to the MALDI-MS spectra obtained from clinical samples in Kazakhstan, we incorporated into our analysis the publicly available MALDI-MS data from South America (3).

### MALDI-MS analysis

Within feasible limits, we closely followed the published methodology for sample preparation, spectra acquisition and preprocessing (3), with only minor modifications as specified. Spectral acquisition was performed on the MicroFlex LT v. 3.4 instrument (Bruker Daltonics, Bremen, Germany) equipped with a pulsed UV laser (N2 laser with 337 nm wavelength, 150 microJ pulse energy, 3 ns pulse width and 20 Hz repetition rate). After thawing at room temperature, samples were spotted onto the steel target plate at 0.5 μl, covered with 0.5 μl of the HCCA matrix (a solution containing α-cyano-4-hydroxycinnamic acid diluted in acetonitrile, 2.5% trifluoroacetic acid and nuclease-free water) and then air dried. The target plate was then loaded into the instrument. Spectra were generated by summing 500 single spectra (10 * 50 shots) in the range between 3 and 20 kDa, operating in positive-ion linear mode using a18-20 kV acceleration voltage, by shooting the laser at random positions on the target spot.

### Spectral preprocessing

Raw MALDI-MS files (Bruker) were uploaded and subsequently preprocessed in R (v. 4.3.0) using MALDIquantForeign and MALDIquant (6). To ensure consistency in peak processing with the original untargeted protocol (3), we used the R scripts generously shared by the authors. Briefly, the spectra were trimmed to a 3–15.5 kDa range, square-root transformed, and smoothened via the Savitzky–Golay method. Baseline correction was done using the TopHat algorithm and intensity normalization was done via total ion current calibration as implemented in MALDIquant. Peak detection was performed using a signal-to-noise ratio of 2 and a halfWindowSize of 10, and the peaks were binned with a tolerance of 0.003. Peak binning was performed in two stages to avoid any additional calibration differences. First, each group spectra were binned separately, and peak filtration was performed, keeping only those peaks that were present in 80% of the spectra of each group. Subsequently, all peaks were binned together. The resulting peak intensity matrix was used for the downstream analyses.

In Analysis I, to assess the models trained on the South American samples from the source study (3), we made slight modifications to the sample preprocessing protocol as follows. To ensure that we are comparing the same 88 peaks, we employed the”reference” method for peak binning using the median values of the spectra and peaks obtained by Nachtigall *et al*. as a reference and eliminated the filtering procedure for each subgroup. In Analysis II, we constructed a *de novo* peak matrix representative of the combined South America and Kazakhstan dataset using the script provided by Nachtigall *et al*.

### Principal component and hierarchical cluster analyses

PCA was performed using R FactoMineR and factoextra packages. The hierarchical cluster analysis was done by first calculating a distance matrix using the Euclidean method and clustering samples via the unweighted paired group with arithmetic mean (UPGMA) method. Dendrograms were generated using ggtree and ggtreeExtra R packages.

### Machine learning and statistical analysis

We implemented a total of seven ML algorithms, six of which were used in the earlier study [5] (DT (Decision Tree - Quinlan’s C5.0 algorithm), KNN (k-Nearest Neighbors), NB (Naive Bayes), RF (Random Forest), SVM-L (Support Vector Machine with linear kernel), SVM-R (Support Vector Machine with radial basis function kernel) plus an additional algorithm XGBoost (eXtreme Gradient Boosting). Analysis 1 was executed by closely following the earlier protocol, with training performed on South American SARS-CoV-2+ and NCARI spectra. Since the training step of analysis II incorporated three sub-groups, i.e. AC samples in addition to the SARS-CoV-2+ and NCARI, the analysis pipeline was modified as outlined below to accommodate this change.

Initially, we split the entire sample into two distinct groups: the training dataset, consisting of 80% of samples, and the test group, which accounted for the remaining 20%. In line with Nachtigall *et al* [5], we conducted a training process using a five-fold (outer) nested repeated (five times) ten-fold (inner) cross-validation with a randomized stratified splitting approach. To optimize the performance of each algorithm, we tested 20 hyperparameters in the inner loop of the cross-validation approach, using a random search method. This process was repeated 20 times to ensure robustness and reliability of the model. We selected the best models based on their area under the curve (AUC) score, which is a common metric for evaluating binary-classification model performance, using the Caret R package. In addition, model performance was assessed using several other classification metrics, including F-measure, recall, accuracy, specificity, sensitivity, and positive and negative predictive values in the yardstick R package; differences across the sub-groups were assessed using the Mann-Whitney U non-parametric test in R.

## Results

The primary objective of the study was to assess the capacity of the MALDI-MS approach to detect SARS-CoV-2 infection within a heterogeneous mix of SARS-CoV-2+, NCARI and AC samples (Table 1).

**Table 1.** Demographic characteristics of participants.

| Characteristic | Overall,<br>N = 252 | SARS-COV-<br>2+ 2021,<br>N = 108 | SARS-COV-2+<br>2022,<br>N = 7 | NCARI,<br>N=98 | AC, N=39 | p-value* |
| --- | --- | --- | --- | --- | --- | --- |
| Age, years,<br>median (IQR) | 38.0<br>(18.0,60.0) | 61.0<br>(48.0,69.0) | 3 (1.0, 37.0) | 8.0 (2.0,<br>35.0) | 34.0 (25.0, 47.0) | <0.001 |
| Male sex, n (%) | 114<br>(45.2%) | 49 (45.4%) | 6 (85,7 %) | 38 (38,8%) | 21 (53.8%) | <0.001 |
| Kazakh ethnicity,<br>n (%) | 116<br>(46%) | 26 (24.1%) | 5 (71,4%) | 61 (62.2%) | 24 (61.5%) | <0.001 |
| Any<br>comorbidities | 104<br>(41.2%) | 72 (66.6%) | 1 (14,2%) | 15 (15.3 %) | 14 (35.9%) | <0.001 |

\* Differences across the groups were assessed using Kruskal-Wallis or Pearson's Chi-squared tests.

Therefore, we performed two independent analyses (Figure 1). In the first analysis, we assessed the performance of the Nachtigall et al. ML pipeline on the combined pool of samples, both from the original study (data collected from three South American countries in 2020) and Kazakhstan (data collected in 2021 and 2022); the ML pipeline in this analysis was trained only on the original South American datasets. In the second analysis, we retrained the ML algorithm, accounting for the spectra contributed by the samples from Kazakhstan and applied this re-trained ML algorithm to the combined pool of samples.

**Fig 1.**
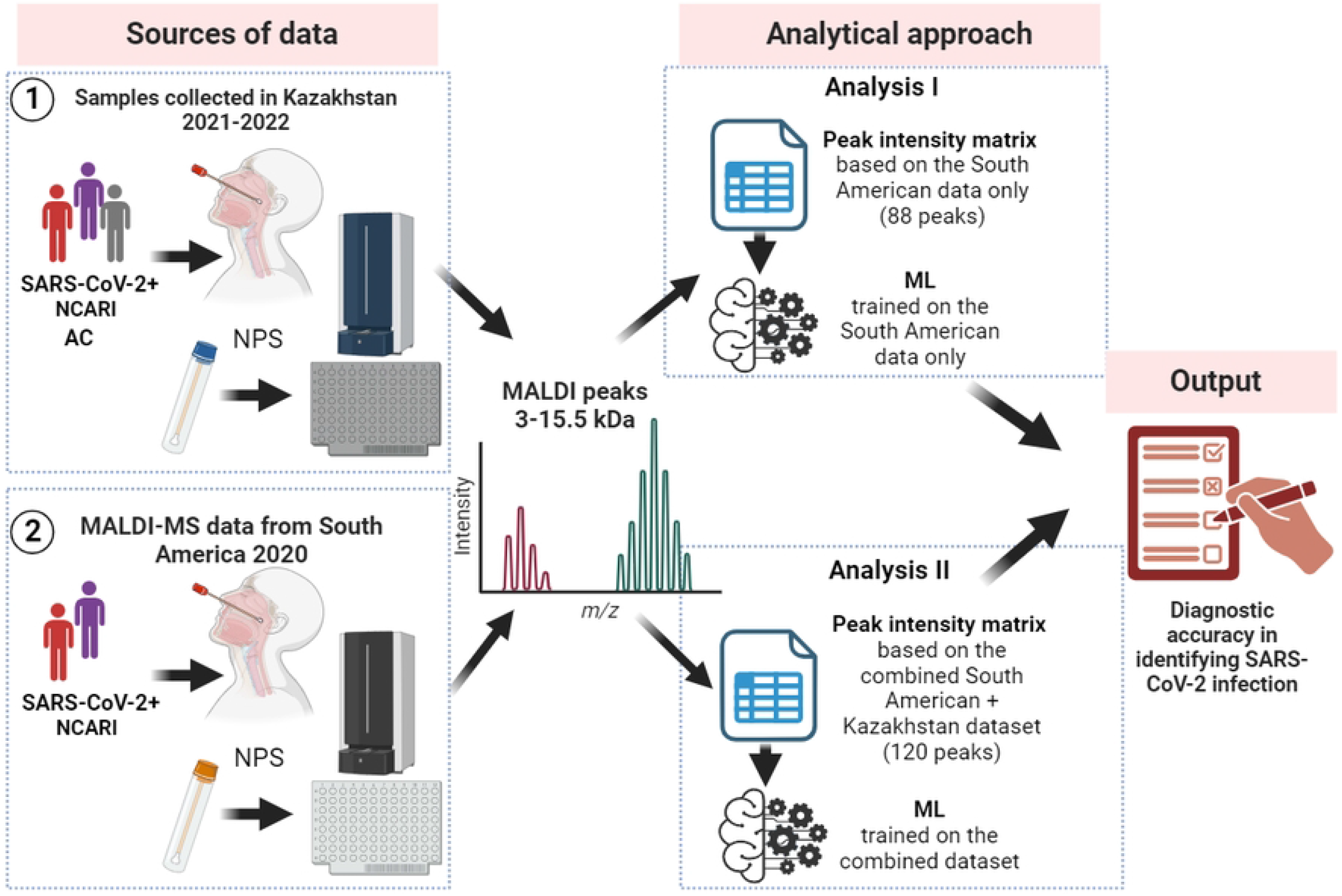
Overall study workflow and description of the analyses. NCARI: non-COVID acute respiratory infections; AC: asymptomatic controls; ML: machine learning

### Analysis I: Applying the”as is” MALDI-MS pipeline to differentiate ARI samples collected in Kazakhstan

To assess how well the original analysis pipeline (3) would differentiate SARS-CoV-2+ samples within the dataset from Kazakhstan, we replicated the steps for i) MALDI-MS peak selection, ii) ML training and iii) ML assessment. Specifically, we focused on the same MALDI-MS peaks that Nachtigall *et al*. (3) used in their analyses (Table S1). These peaks were derived using a six-step spectra processing workflow including spectra transformation and smoothing, baseline removing, spectra calibration, peak detection, and peak processing.

We then constructed a peak intensity matrix on the 88 peaks, identical to that used by Nachtigall and colleagues (3), for the downstream analysis of a combined dataset incorporating both the South American (Table S1) and Kazakhstan samples (Figure 2 and Table S2).

**Fig 2.**
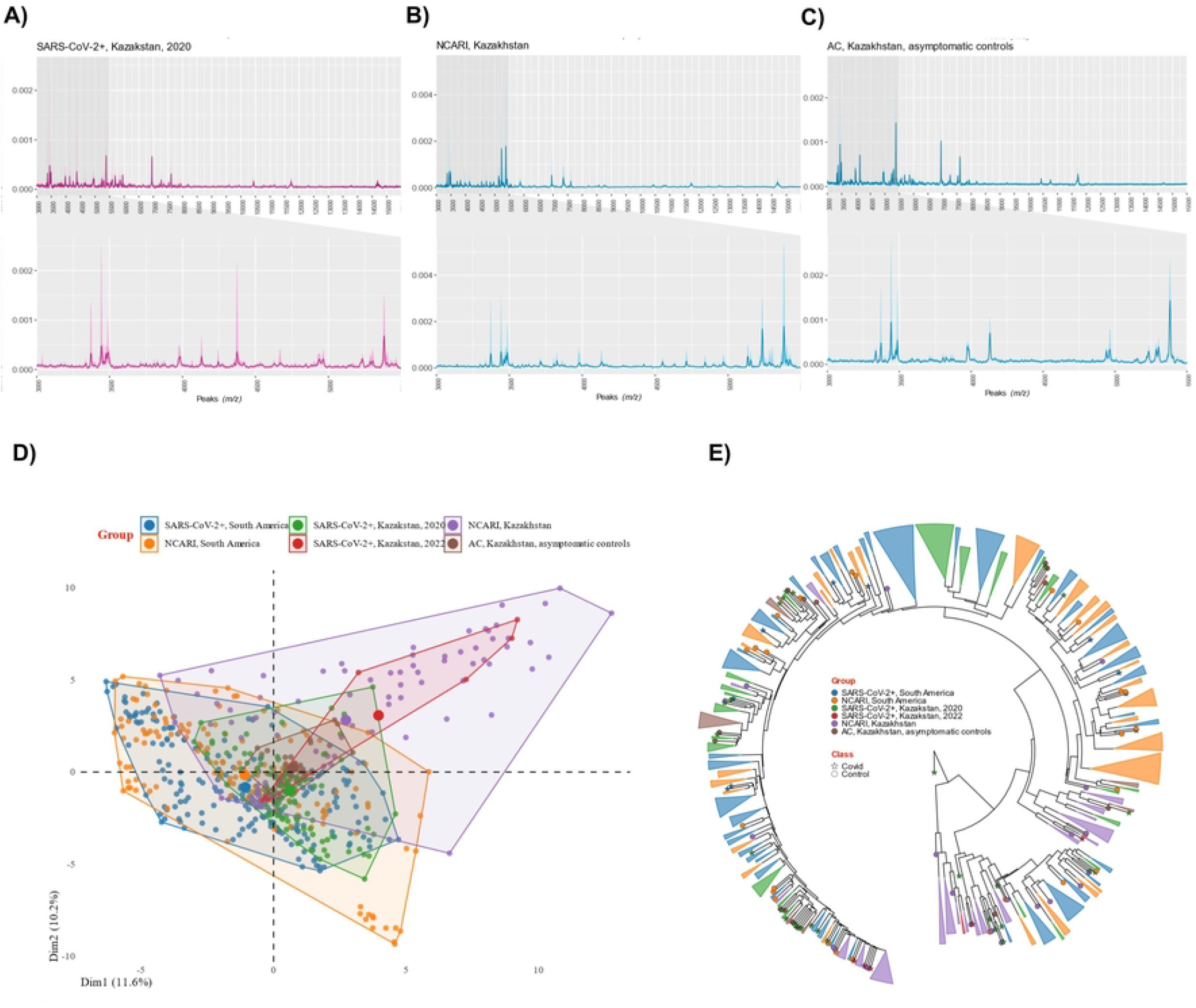
MALDI-MS peak data generated using nasopharyngeal swabs and processed following the MALDI-MS/ML pipeline developed by Nachtigall and colleagues (3). A-C. representative MALDI-MS spectra from symptomatic SARS-CoV-2+ (A), symptomatic non-SARS-CoV-2 (B) and a healthy control sample from Kazakhstan (C). The central line indicates median value of the spectra, while the shaded region on either side represents the interquartile interval. Insets depict a range from 3000 to 5500 m/z encompassing 70% (62/88) of the identified peaks. d. PCA of the combined dataset incorporating MALDI-MS data both from Kazakhstan and South America (2020 SARS-CoV+ and symptomatic SARS-CoV-2-negative)(3).

We next explored the selected peaks across the comparison groups by reducing the multidimensionality using principal component analyses and dendrograms. Like Nachtigall et al, we did not detect any obvious clustering by sub-group, emphasizing the need for a more sensitive approach to discern subtle differences in the highly multidimensional MALDI-MS peak data (Figure 2D and 2E, Figures S2-S5). Hence, we then applied to our combined Kazakhstan-South America MALDI-MS peak dataset the original Nachtigall *et al*. ML algorithm trained on the original South American samples (3).

In keeping with earlier results (3), when tested the South American samples alone, SVM-R provided the highest ROC AUC, although other models had similarly high-performance characteristics (Table S3 and Figure 3A-B) for classifying cases of SARS-CoV-2 and non-SARS-CoV-2.

**Fig 3.**
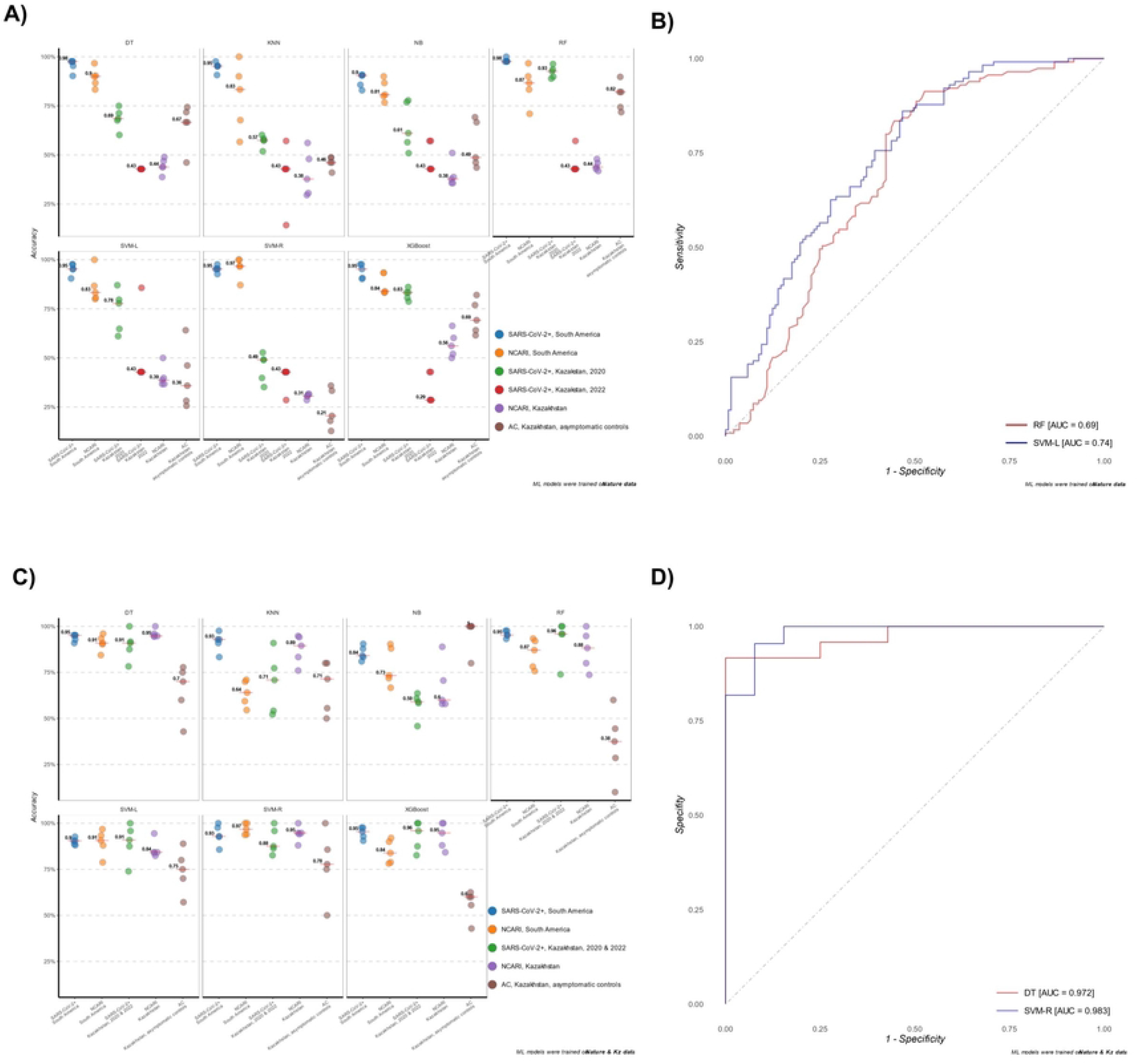
Classification accuracy of the MALDI-ML algorithms assessed on the data from Kazakhstan and South America. A) Accuracy metrics for each of the seven ML models trained on the South American MALDI-MS data (Analysis I in the current study) for the differentiation of study sub-groups. B) ROC curves of the top-performing RF and SVM-L algorithms (Analysis I). C) Accuracy metrics for each of the seven ML models trained on the combined South America-Kazakhstan dataset (Analysis II in the current study) for the differentiation of study sub-groups. D) ROC curves for the top-performing SVM-R and DT algorithms (Analysis II).

Subsequently, we assessed the performance of the same ML algorithms on samples from Kazakhstan. Here, we observed a broad variation in the ability of the ML models to discern SARS-CoV-2+ samples. RF had the highest percentage of correctly identified 2020-SARS-CoV-2+ samples (91%) (Figure 3A and Table S4, Figure S6). Notably, the accuracy for 2021 SARS-CoV-2 was <60% for all models, similar to the accuracy for identifying NCARI. RF discerned AC with an accuracy of 68%, the highest of all models for this sub-group.

### Analysis II: Applying the re-trained MALDI/MS-ML to differentiate ARI

To ensure that we include all relevant MALDI-MS signature peaks representative of all sub-groups, we performed peak selection on the entire pool of samples containing samples from both Kazakhstan and South America (n=615). A total of 120 peaks were identified and a peak intensity matrix was constructed (Table S5). As in Analysis I, PCA and dendrograms did not show any visually apparent clustering of sub-groups (Figures S7-S10). We then proceeded to train ML models on the combined pool consisting of the 120 peaks, of which 53 overlapped with the original 88 peaks.

Due to the small sample size of the 2022 subset, the SARS-CoV-2 2021 and 2022 subsets were combined prior to testing the model performance. We then assessed the performance of the trained ML algorithm on the South America-Kazakhstan dataset. All models demonstrated similarly high-performance characteristics in differentiating SARS-CoV-2+ samples. SVM-R and DT slightly outperformed the other five models in discerning SARS-CoV-2 infection from both NCARI and AC with ROC AUC values of 0.983 [0.958, 0.987] and 0.972 [0.966, 0.979], respectively (Figures 3C-D and Table S4). SVM-R, in particular, differentiated the Kazakhstan SARS-CoV-2+, NCARI, and AC subjects with an accuracy of 88.0, 95.0 and 78.0%, respectively (Figure 3C). Both SVM-R and DT were also highly accurate at differentiating NCARI and AC sub-groups (Table S4).

## Discussion

Here we aimed to assess the feasibility of deploying MALDI-MS and ML in a clinical lab to differentiate SARS-CoV-2 from other ARI, particularly in the context of minimal specialized sample preparation. Our initial application of the original MALDI-MS/ML pipeline, trained on South American samples (3), demonstrated reduced efficiency in identifying samples from Kazakhstan. Re-training the ML models to incorporate MALDI peak information from a diverse pool of Kazakhstan samples, including SARS-CoV-2+, NCARI subjects, and asymptomatic controls, led to a significant improvement in detection accuracy. Taken as a proof-of-concept, our results support the utility of MALDI-MS/ML, especially in the early phases of respiratory endemics/pandemics, when limited knowledge is available on the infectious pathogen’s identity and in low-resource environments, where alternative methods may yet be unavailable.

Our replication studies underscore the importance of considering geographical and population-specific variations in the application of MALDI-MS/ML. The observed differences in the performance of the original pipeline trained on South American samples may be attributed to the inherent complexity of NPS, which contains a mixture of host proteins and diverse microbial species (7,8). The sensitivity and specificity of MALDI-MS/ML may also be affected by variability in immune response to different viral loads and the presence of co-infections (9,10). These challenges emphasize the need for careful evaluation and calibration in the application of MALDI-MS/ML.

Our study has several limitations. The lack of specialized sample preparation, although advantageous for low-resource settings, may introduce variability and noise into the data, a concern raised by other authors (9,10). Due to a relatively small sample size of the NCARI group, we did not further pursue stratification of this group by the causative agents identified via multiplex PCR. The utility of MALDI-MS/ML in differentiating various NCARI would be important to examine in the context of the changing post-pandemic ARI landscape (11). Temporal variation, spanning samples collected over two years (2020-2022), might have contributed to a high heterogeneity of our results. The differences across groups regarding the basic demographics may also have a confounding effect on the results. Further validation of the method in a broader clinical context would be necessary to fully assess the potential for real-world application.

In conclusion, our study provides valuable insights into the potential of MALDI-MS as an accessible laboratory-based diagnostic tool for ARI. While promising, the implementation of MALDI-MS/ML in real clinical lab settings will require further optimization, validation, and continuous adaptation to the evolving epidemiological landscape. Further research is needed to explore the specific components of MALDI-MS spectra that are most informative for differentiating various ARI. Such investigations will contribute to the ongoing refinement of this promising diagnostic tool.

## Data Availability

All raw data and R code are available through Github (https://github.com/dimbage/ML_MALDI-TOF_SARS-CoV-2).

## Declarations

### Ethics approval and consent to participate

All study procedures were approved by the Research Ethics Board of Karaganda Medical University under Protocol 12 (approved 45) from 06.04.2020. Written informed consent was obtained from all participants.

### Consent for publication

Authors provide consent for the publication of the manuscript detailed above, including any accompanying images or data contained within the manuscript.

## Acknowledgements

We thank the study participants and clinic staff. We are grateful to Professor Leonardo Santos for sharing the R scripts and associated data from their original study.

## Supplementary information

All supplementary information can be found in the Appendix.

